# Trial participants are frequently excluded based on their symptoms rather than their condition: A systematic review of Cochrane reviews and their component trials

**DOI:** 10.1101/2023.04.21.23288922

**Authors:** Katie Stocking, Andrew Watson, Jamie J Kirkham, Jack Wilkinson, Andy Vail

**Affiliations:** Centre for Biostatistics, Faculty of Biology and Health Sciences, University of Manchester, Manchester, UK; Department of Obstetrics and Gynaecology, Tameside & Glossop Acute Services NHS Trust, Ashton-Under-Lyne, UK

**Keywords:** Research waste, Randomised Clinical Trial, Cochrane Review, Gynaecology, Outcome selection

## Abstract

**Objectives:** **I**dentify strategies used in the design of recent randomised controlled trials (RCTs) and their associated Cochrane reviews where patients with the same gynaecological condition present with different symptoms.

**Study Design and Setting:** We searched the Cochrane library (February 2022) for reviews in polycystic ovarian syndrome (PCOS) and endometriosis. Reviews were included if the intervention was intended to treat all condition-specific symptoms. We restricted to trials published since 2012 to consider ‘current’ approaches. For each trial we recorded the number of potentially eligible participants excluded as a direct result of the chosen strategy. For each review we recorded the numbers of RCTs and participants excluded unnecessarily.

**Results:** There were 89 distinct PCOS trials in 13 reviews, and 13 Endometriosis trials in 11 reviews. Most trials restricted their eligibility to participants with specific symptoms (55% PCOS, 46% endometriosis). The second most common strategy was to measure and analyse clinical outcomes that were not relevant to all participants (38% PCOS, 31% endometriosis). Reviews excluded 27% of trials based just on outcome data.

**Conclusions:** Current gynaecological research is inefficient. Most trials either exclude patients who could benefit from treatment or measure outcomes not relevant to all participants.

**Registration:** PROSPERO (CRD42022334776)

**What is new?:** *Key findings:*

- Over a quarter of Cochrane reviews included in this review excluded trials based on the outcomes reported.
- Typically, recent randomised controlled trials in Polycystic Ovarian Syndrome and Endometriosis trials either exclude patients who could potentially benefit from the treatment given, or measure outcomes of no relevance to some participants.

*What this adds to what is known?:*

- Strategies developed that are employed in the design and measurement of outcomes in gynaecological trials.
- There are multiple sources of waste in the current gynaecological research landscape. The population of patients available is under-utilised by excluding patients based on the outcomes measured, or alternatively, researchers are measuring outcomes in patients who do not experience the associated symptom(s).

*What is the implication and what should change now?:*

- Gynaecological patients experience heterogeneity in their symptoms and therefore it is crucial to employ appropriate outcome measures in order to reduce research waste. Cochrane Reviews should include all trials which report outcomes that are relevant to the population of interest if the intervention under investigation is deemed to plausibly treat the associated symptom(s).

## 1. Introduction

The principal role of a randomised controlled trial (RCT) is to evaluate whether a medical intervention is safe and effective. In order for this to happen, it is imperative that researchers measure outcomes which are both appropriate and relevant to the population of interest. Although randomised controlled trials remain the gold standard tool for treatment evaluation, many, through poor design, contribute to the overwhelming problem of waste in research [1-4]. In the *Lancet* collection of papers on waste in medical research, it was estimated that $240 billion of annual research expenditure is wasted [3, 5-8]. It is indeed true that much work is being done to reduce this figure, however, there is still much room for improvement [9, 10]. Inefficient studies that fail to address questions that matter to both patients and stakeholders emphasise the importance that we need to do less, but better, research [11].

Often, in the case of gynaecological conditions such as Polycystic Ovarian Syndrome (PCOS) and Endometriosis, patients require different things from their care, at different stages of their lifetime [12-15]. Not all patients with the same diagnosis will experience all of the associated complications, and, although their most bothersome symptom might differ, it is common to receive the same treatment. If we take PCOS, for example, the Rotterdam criteria are the most widely used classification for diagnosis and proposes that PCOS is present if the patient has at least two of the three characteristics: oligo- and/or anovulation, clinical and/or biochemical hyperandrogenism, and polycystic ovaries on ultrasound [16]. These criteria for the diagnosis of PCOS in itself has consequences, as by definition, not all patients with PCOS have all the possible manifestations of the disorder and therefore do not experience the same symptoms and health risk factors [12]. For endometriosis, patients may have symptoms dominated by pelvic pain, infertility, or both. In the post-reproductive era the reduction in quality-of-life from menstrual dysfunction may predominate. In the likely scenario that potential trial participants have no, or little, symptoms in common, it presents the problem of how researchers select relevant, patient-important outcomes and design RCTs that are not wasteful.

One of the most notable inclusions to the movement to reduce waste in research is the development of core outcome sets (COS). The Core Outcome Measures in Effectiveness Trials (COMET) Initiative encourages the application of agreed standardised sets of outcomes. These outcome sets represent the minimum that should be measured and reported in clinical trials in specific clinical areas [17-19].

Recently developed core outcome sets in both Polycystic Ovarian Syndrome (PCOS) and endometriosis undoubtedly violate the concept of COS, as authors in both instances concluded that not all outcomes could be reported in all trials [20, 21]. This is demonstrative of the fact that in the field of gynaecology, it is not one-size-fits-all.

This prompts the question of how we should design trials so as to incorporate this heterogeneity, as well as the implications for systematic reviews and meta-analysis. Due to the multifactorial nature of a patient’s symptoms, the design and recruitment of gynaecological trials is challenging and is approached in different ways. We conducted a systematic review to investigate how diverse symptoms and patient populations are currently handled in gynaecological trials, in which the intervention could plausibly be used to treat all symptoms related to the diagnosis. We aimed to identify the methodological strategies applied within randomised controlled trials in Cochrane Reviews, where an intervention is hypothesised to have potential benefit for patients with the index condition.

## 2. Methods

The study design was a systematic review with descriptive statistics. Our overall approach was to identify systematic reviews in the conditions of interest and to examine the characteristics and methodological practice of their included and excluded trials. Protocol registration was with the Prospective Register of Systematic Reviews (registration number: CRD42022334776). Full details of methods are given there but summarised below.

In February 2022 searches were undertaken to identify systematic reviews contained in the Cochrane Library on interventions for PCOS or endometriosis. We considered trials list under both ‘included’ and ‘excluded’ categories.

### 2.1 Study inclusion criteria

Decisions regarding eligibility were made by discussion with AW, a consultant in gynaecology.

Cochrane intervention reviews and RCTs were eligible for inclusion from 2012 onwards to give an overview of current practice. Cochrane reviews were included only if the intervention under investigation was intended to treat the underlying condition, and would therefore plausibly treat all condition-specific symptoms. For example, in vitro fertilisation would not be an eligible treatment, as it would be used only for fertility outcomes.

### 2.2 Data extraction

Two reviewers (KS and AW) screened all titles and abstracts against the inclusion criteria. Any disagreements were resolved through discussion.

Outcomes categories were pre-specified (appendix), along with whether each outcome was primary, secondary, or unspecified. This information was also extracted for each of the trials, along with the setting, intervention type, funding source, design, size (number of participants randomised), number of participants excluded for symptom/outcome-related reasons and the number of participants contributing to the primary outcome.

Seven different strategies were anticipated for the RCTs, as demonstrated in Figure 1. We developed these strategies as part of an iterative process. As a pilot, we developed some potential strategies we had seen during our time as researchers, assessed several trials and determined if the strategy used by each trial was different to those we anticipated. We then discussed and re-evaluated until no new strategies were found. We note that this may not be an exhaustive list, and we considered whether any additional strategies not included in our list were used. Each trial was categorised according to the strategy used, in the order listed, i.e. if the trial did not fit the criteria to be the first strategy, it was considered for the next, and so on.

**Figure 1.**
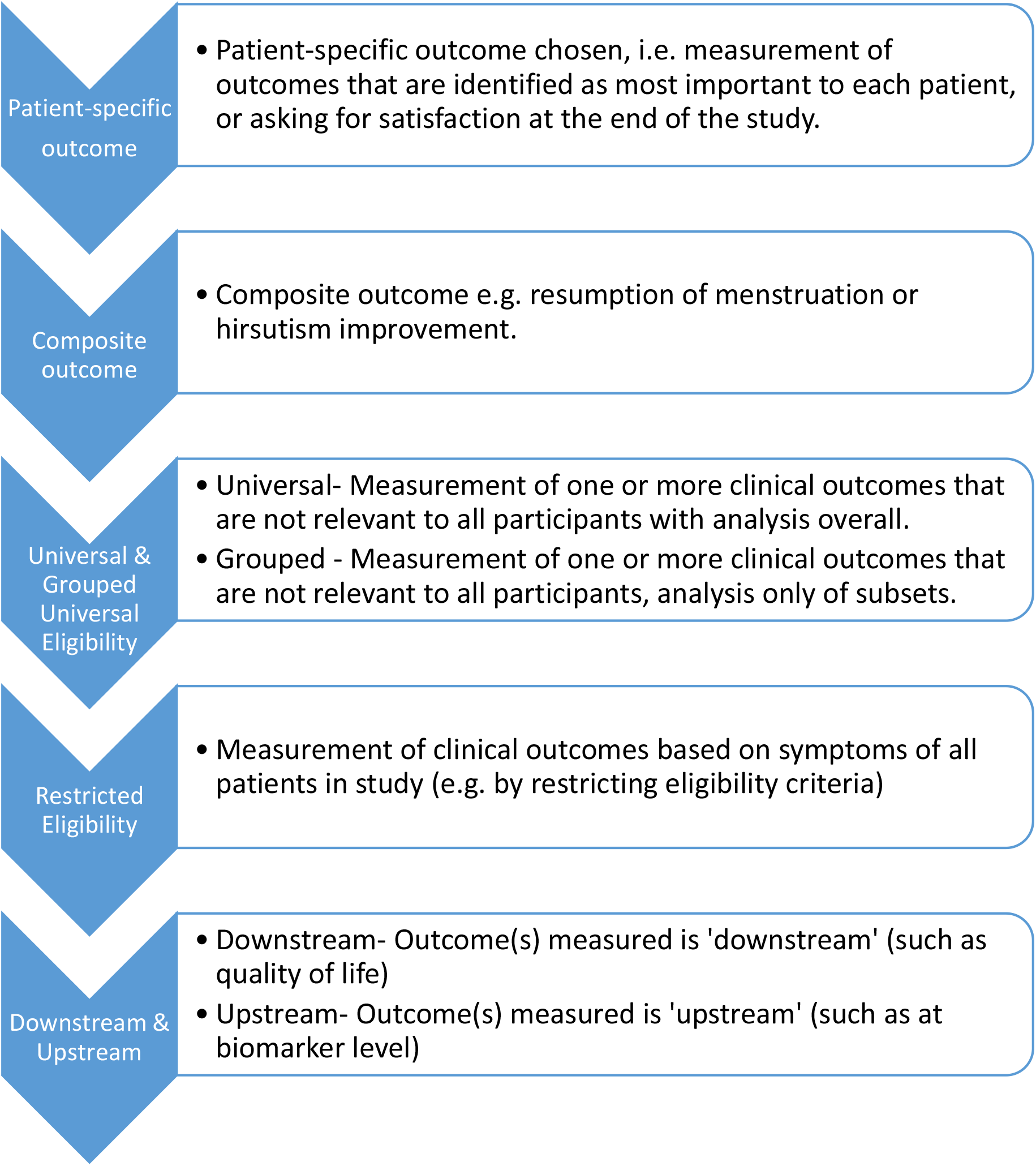
Strategies for randomised controlled trials in gynaecology.

For ease of context and reference, we named the strategies and will refer to them as such throughout:

- Participant-specific Outcome Strategy - patient-specific primary outcome was chosen, for example patient satisfaction or success.
- Composite Outcome Strategy - composite outcome, for example the resumption of menstruation or hirsutism improvement.
- Universal Eligibility Strategy - measured outcomes regardless of their relevance to the patients in the study, e.g. a study which did not restrict to patients experiencing amenorrhea (a lack of menstruation) but measured resumption of menstrual period as an outcome in every patient.
- Grouped Universal Eligibility Strategy would be trials designed the same way as the Universal Eligibility Strategy, but the statistical analysis would be restricted to those who experienced amenorrhea at baseline, as a subset analysis.
- Restricted Eligibility Strategy was allocated where a trial measured only clinical outcomes based on the symptoms of all patients in the study, i.e. a fertility trial, interested only in fertility-based outcomes, where the inclusion criteria is patients experiencing subfertility.
- Downstream Eligibility strategy was studies measuring only quality-of-life outcomes.
- Upstream Eligibility strategy was studies reporting no clinical outcomes, only biomarker outcomes.

For each trial, where available, we recorded the number of potentially eligible participants excluded as a direct result of the chosen strategy relative to the achieved sample size. That is, how much larger could the recruitment have been without an outcome-defined restriction on eligibility criteria. Where available, the number of potentially eligible participants were taken from the CONSORT flow chart. This was taken to be the number of participants that were found to be ineligible for outcome-related reasons pre-randomisation. Similarly, for each Cochrane review we recorded the numbers of identified RCTs excluded from consideration for reasons such as not reporting review-specific outcomes. To determine which trials were excluded from each Cochrane review and the reason for exclusion, we used the ‘Characteristics of excluded studies’ section of the review. We selected all excluded trials for assessment where the reason for exclusion was related to the population of patients, outcome reporting or selection.m

### 2.3 Study quality assessment and data analysis

We accepted the published risk of bias assessment for the included studies. For the studies that were excluded from the Cochrane review, and therefore have no associated risk of bias assessment, these were independently assessed using Cochrane’s risk of bias tool as a surrogate for their quality.

Descriptive analyses were undertaken. All data synthesis was exploratory and included the calculation of a mean number of exclusions.

We aimed to analyse whether calendar year of publication and study-level factors (e.g. sample size, nature of intervention, study quality assessed by risk of bias) were associated with inefficiency, using Fisher’s Exact test as an exploratory analysis. For this, date of publication was divided into pre-2017 and 2017 or after; nature of intervention was medical, surgical or other; risk of bias was judged as either in the primary analysis or not if selection bias was deemed low using Cochrane Gynaecology and Fertility’s guidance [22] and strategy was compared as Universal Eligibility, Restricted Eligibility or Other strategy.

## 3. Results

### 3.1 Study selection and characteristics

For PCOS, there were 31 Cochrane reviews screened at the title and abstract stage, of which 13, containing 239 trials (included those that were excluded from the associated Cochrane review), met the inclusion criteria. There were 136 trials which were excluded from this review for reasons relating to design and access, information can be found in the PRISMA (Figure A1). Duplication occurred in two ways, the first, where the same trial appeared in multiple Cochrane reviews. We removed these duplicate trials, including only the first time, chronologically, that the trial appears in a review (2 trials appeared three times, 7 appeared twice). The second form of duplication occurred on 3 instances where publications had re-analysed original trial data, reporting different outcomes. In this case, all data were collected and considered as one trial. Therefore, a total of 89 trials in 13 Cochrane reviews contributed to the findings of this review.

For endometriosis, 32 Cochrane reviews were screened at the title and abstract stage with 11, containing 19 trials, meeting the inclusion criteria. Six trials were excluded as they were abstract only (n=3), inaccessible (n=1) or had no locatable publication (n=2), therefore, a total of 13 trials in 11 Cochrane reviews were included in our systematic review (Figure A2). The trial characteristics are summarised in Table 1. Most commonly, PCOS patients were recruited from obstetrics and gynaecology clinics (40%) and fertility clinics (35%), with very few research teams recruiting from the community (4%). The majority of endometriosis patients were recruited from obstetrics and gynaecology clinics (85%).

**Table 1.**
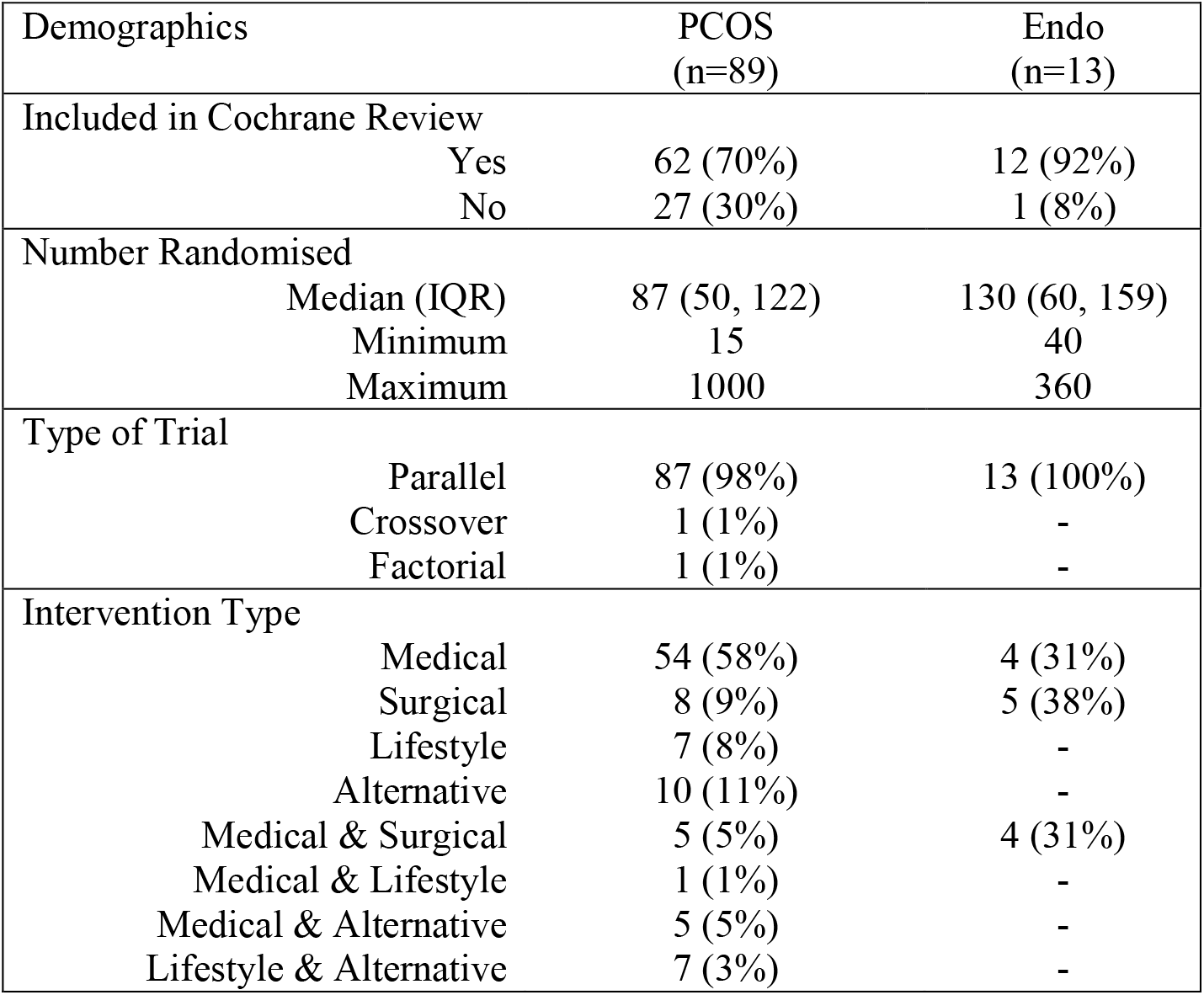
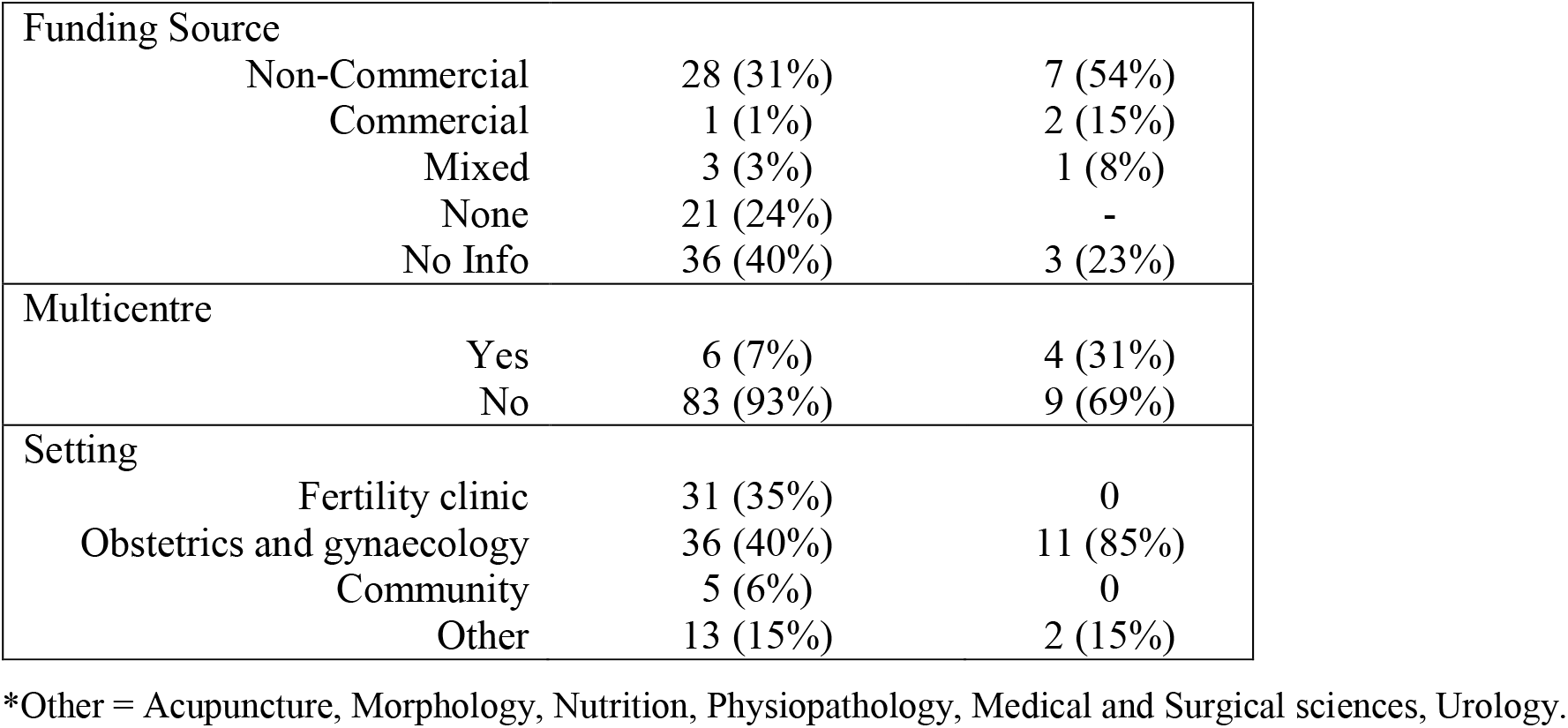
Characteristics of RCTs identified in PCOS and endometriosis

### 3.2 Trial strategies

Only 7 trials included in this review reported the number of participants excluded pre-randomisation, for outcome-based reasons. However, these 7 trials excluded a total of 990 participants (median 16, IQR: 4-229, minimum: 3, maximum: 704). To give context to this, the total sample size accrued by these 7 trials was 2744 (median 172, IQR: 46-750, minimum: 45, maximum: 1000).

The most common strategy used by researchers in PCOS and endometriosis was Restricted Eligibility (55% and 46%, respectively). In PCOS, we found that over half of the trials that used Restricted Eligibility focused solely on fertility-based outcomes (59%), demonstrated in Table 2. For endometriosis there was more variation in the outcomes reported, with a third (33%) of Restricted Eligibility studies reporting pain only outcomes and a third (33%) reporting fertility only outcomes.

**Table 2.** Proportion of outcome combinations in each strategy.

| Outcome Combination | Strategy |  |  |  |  |  |  |  |
| --- | --- | --- | --- | --- | --- | --- | --- | --- |
|  | Universal Eligibility |  | Grouped Universal Eligibility |  | Restricted Eligibility |  | Upstream Outcome |  |
|  | PCOS (n=34) | Endo (n=4) | PCOS (n=1) | Endo (n=2) | PCOS (n=49) | Endo (n=6) | PCOS (n=5) | Endo (n=1) |
| Fertility only | 0 | 0 | 0 | 0 | 29 (59%) | 2 (33%) | 0 | 0 |
| Pain only | 0 | 0 | 0 | 0 | 0 | 2 (33%) | 0 | 0 |
| Fertility + QoL + Other clinical outcome(s) | 6 (18%) | 0 | 0 | 0 | 0 | 0 | 0 | 0 |
| Fertility + Other clinical outcome(s)* | 10 (29%) | 2 (50%) | 1 (100%) | 0 | 13 (26%) | 0 | 0 | 0 |
| QoL + Other clinical outcome(s) | 4 (12%) | 0 | 0 | 2 (100%) | 0 | 1 (17%) | 0 | 0 |
| Clinical outcome(s) + Treatment satisfaction | 0 | 2 (50%) | 0 | 0 | 0 | 1 (17%) | 0 | 0 |
| Other clinical outcome combinations | 14 (41%) | 0 | 0 | 0 | 7 (14%) | 0 | 0 | 0 |
| Biomarkers only | 0 | 0 | 0 | 0 | 0 | 0 | 5 (100%) | 1 (100%) |

Percentage given is within condition. \*fertility + pain for endometriosis

Universal Eligibility was the second most used, with 38% of PCOS trials using this approach. Of these trials, the majority measured combinations of outcomes, most commonly choosing to measure multiple clinical outcome combinations (41%) or fertility and other clinical outcomes (29%). There were 31% of endometriosis trials that used this strategy. Similarly to the PCOS trials, they measured combinations of outcomes: fertility and pain (50%) and patient important plus other outcomes (50%)

No PCOS trials and only one endometriosis trial that employed the Universal Eligibility strategy made note of the numbers of patients experiencing the primary outcome at baseline. Therefore, we were unable to calculate the ratio of patients not experiencing the primary outcome of interest in relation to the sample size randomised.

Grouped Universal Eligibility was used in 15% of identified endometriosis trials, and only 1% in PCOS trials. The Upstream Outcome strategy was used in 8% of endometriosis trials, and 6% of PCOS trials. There were no instances where the Patient-specific Outcome, Composite Outcome or Downstream Outcome strategies were used.

There was no significant difference in the strategies used in pre-2017 compared to 2017 and after in PCOS (p=0.76) or endometriosis (p=0.63). Interventions tested were also not significantly different between trial strategies in PCOS (p=0.05) or endometriosis (p=0.27). When comparing the studies which could plausibly have been included in a primary analysis, using the RoB assessment, only 5 out of the 102 possible PCOS and endometriosis trials were classified as low risk.

### 3.3 Review-level findings

Of the 27 trials excluded from the Cochrane reviews in PCOS, most were excluded as the authors of the review were interested in fertility outcomes: 74% (n=20) because they were non-fertility studies, 15% (n=4) were PCOS with or without infertility. The remaining trials were excluded for having no outcomes of interest (n=2, 7%) and fertility outcomes only (n=1, 4%). These 27 trials had a median number of 60 people randomised, (IQR: 45-88, minimum: 26, maximum: 233). Similarly, the trial excluded from the endometriosis reviews was on the basis of not reporting the review’s outcome of interest. Overall, the exclusion of trials on the basis of the outcomes they reported totalled 27% of available RCTs (28/102).

## 4. Discussion

We conducted our systematic review to examine the current strategies used for the design of and recruitment to randomised controlled trials, by researchers. We focused on PCOS and endometriosis as exemplar conditions due to the heterogeneity of their symptoms, however, we anticipate the findings to be applicable to gynaecological research in general, and in other conditions where patients experience varying symptoms.

Systematic reviews are considered the gold standard of evidence for decision-making, used to collate, critique and summarise evidence. However, we found that the Cochrane reviews included in this review, excluded many trials, based on the outcomes they reported. Whilst at first glance, choosing a research question, and selecting trials that report outcomes relating to that research question does not appear to be erroneous, there are issues with this. Our review considered only Cochrane reviews of interventions which could, plausibly, be used regardless of symptoms. Where authors exclude trials based on their outcomes, they are reducing the number of patients with data available to add to the evidence base. Dwan and colleagues have previously discussed the prevalence and impact of excluding studies from reviews on the basis of the relevance of outcome data, with further works observing that doing so leads to potentially biasing the conclusions of systematic reviews, along with waste in production and reporting of research [23, 24]. Therefore, it would be advantageous for reviews to consider trials regardless of reported outcomes, and instead consider a multitude of symptoms and outcomes that could plausibly be treated with the intervention under investigation. This could in turn reduce the number of reviews needed, and hence waste from duplication of effort.

In order to tackle the demonstrated waste in gynaecological trials we need to focus first on improving the quality of the research questions, ensuring they matter to patients and clinicians. Three of the strategies we anticipated: Participant-specific Outcome, Composite Outcome, and Downstream Outcome were not being utilised and require consideration. It is likely that these strategies are not currently perceived as attractive to use as those that are more common. Participant-specific outcomes, such as measuring a patient’s most bothersome symptom or allowing a patient to set their own personalised goals, are relatively novel ideas, and although they have previously been used in gynaecological trials, further consideration is needed for their statistical advantages [25, 26].

Composite outcomes allow research to address more than one aspect of a patient’s health status, but their use is widely debated, with interpretation difficult [27-30]. Similarly, although a well-established instrument for providing evidence of an individual’s physical, emotional and social health, interpretation of treatment effect can also be difficult when using a quality-of-life measurement. Cox *et al* suggests that “in practice the main difficulty is likely to be in separating the real treatment-by-individual interaction from noise” [31].

In the current research landscape, we are typically seeing two main strategies employed in gynaecological trials, which we refer to as Universal Eligibility and Restricted Eligibility. In the first, outcomes measured are of relevance to everybody in the trial. This means patients are excluded due to stringent inclusion criteria and are unable to be involved in a trial of treatment that may be of clinical benefit to them. Secondly and conversely, some trials include participants which are not experiencing the symptom at baseline, resulting in the inability to provide useful information, as they cannot contribute to the outcome of interest. For example, researchers record BMI as an outcome for patients not overweight at baseline. In April 2022, the Royal College of Obstetricians and Gynaecology (RCOG) reported that gynaecological care waiting lists in England had grown the most substantially compared to any other medical specialty, seeing an over 60% increase on pre-pandemic levels [32].

There is no shortage of people seeking and requiring gynaecological treatment. Both of the strategies this review identified as most prevalent in gynaecological research do not utilise the potential population and are inefficient, often leading to research that lacks in impact and results in waste.

## 5. Conclusions

As a minimum, participants whose symptoms could potentially benefit from any specific treatment should have a chance to receive said treatment. This research identified multiple sources of waste in the current gynaecological research landscape. We have shown that the population of patients available is often under-utilised by excluding patients based on the outcomes measured, or alternatively, researchers are measuring outcomes in patients who do not experience the associated symptom(s).

Gynaecological patients experience heterogeneity in their symptoms and therefore it is crucial to employ patient-specific outcome measures. Not only would this reduce research waste from a budget and resource perspective, it would also, most importantly, alleviate patient burden.

## Data Availability

All data produced in the present study are available upon reasonable request to the authors

## Acknowledgements

The authors thank the Public Patient Involvement group that have given their time to help support this research and shape the strategies developed.

## Authors’ contributions

Katie Stocking: Conceptualization; Data curation; Formal analysis; Methodology; Writing – Original draft preparation. Andrew Watson: Conceptualization; Data curation; Validation; Writing – Reviewing and editing. Jamie J Kirkham: Conceptualization; Methodology; Supervision; Writing – Reviewing and editing. Jack Wilkinson: Methodology; Supervision; Writing – Reviewing and editing Andy Vail: Conceptualization; Methodology; Supervision; Writing – Reviewing and editing.

