## Appendices for "Trial participants are frequently excluded based on their symptoms rather than their condition: A systematic review of Cochrane reviews and their component trials"

**Appendix**

**PCOS outcome groupings**

1. Fertility effectiveness (e.g. live birth, ongoing pregnancy, clinical pregnancy), Fertility adverse events (e.g. OHSS, Multiple pregnancy, miscarriage) and Ovulation (including ovulation rate, resumption of ovulation)
2. Resumption of menstrual regularity (e.g. oligomenorrhea)
3. Body composition (e.g. body mass index (BMI) ($kg/m^2$), waist circumference (cm), waist-hip ratio (WHR))
4. Assessment of Hirsutism
5. Assessment of Acne
6. Mental Health
7. Quality of Life and Treatment Satisfaction
8. Any biomarker (e.g. androgen levels, metabolic biochemical factors, diabetic markers, fertility markers, etc)

**Endo outcome groupings**

1. Fertility effectiveness (e.g. live birth, ongoing pregnancy, clinical pregnancy), Fertility adverse events (e.g. OHSS, Multiple pregnancy, miscarriage) and Ovulation (including ovulation rate, resumption of ovulation)
2. Resumption of menstrual regularity (e.g. oligomenorrhea)
3. Pain with periods/dysmenorrhea
4. Painful sex / dyspareunia
5. Intestinal symptoms (e.g. diarrhoea, constipation)
6. Urinary symptoms (e.g. retention, neurogenic dysfunction)
7. Pelvic pain
8. Overall pain
9. Mental health
10. Biomarkers
11. Quality of life and Treatment Satisfaction
12. Most bothersome symptom improvement

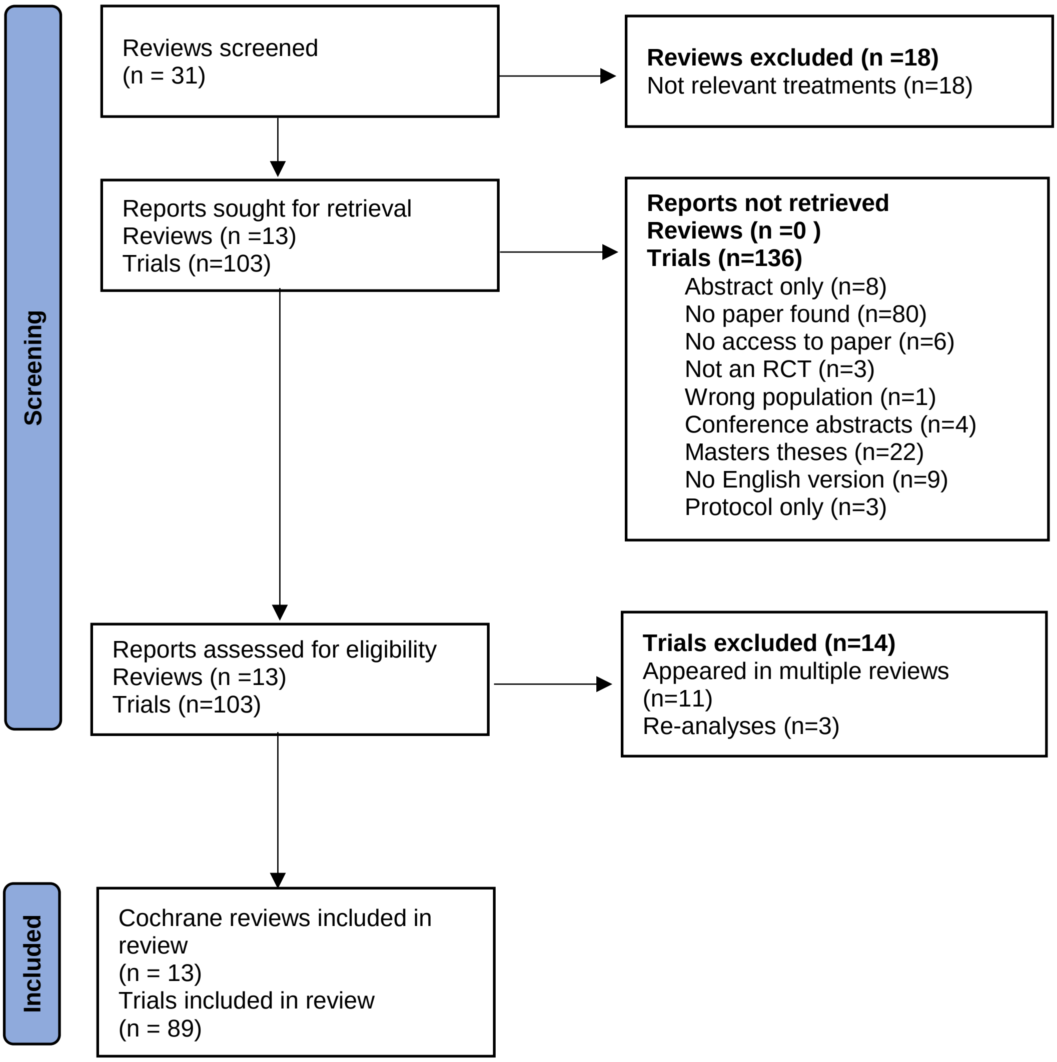


*Figure A1 - PRISMA for Polycystic Ovarian Syndrome*

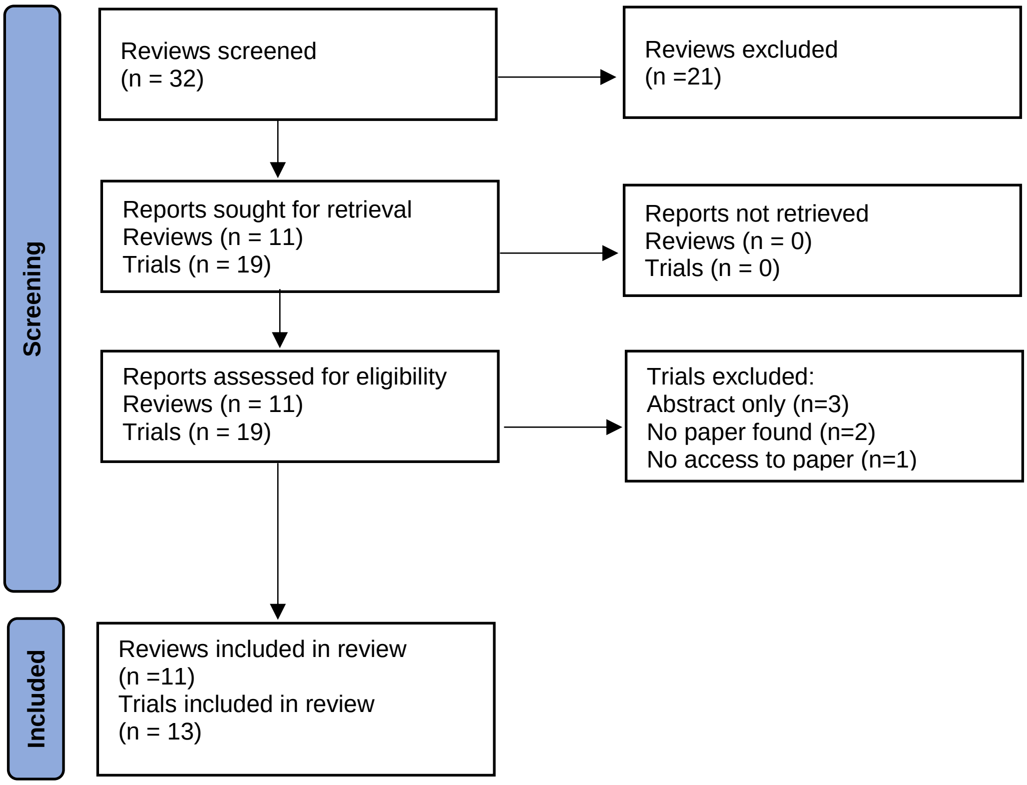


*Figure A2 - PRISMA for Endometriosis*
